# Ethnic and Socioeconomic Inequalities in Health and Social Care Utilisation Among People with Dementia: A Population-Based Study

**DOI:** 10.64898/2026.06.04.26354916

**Authors:** Georgina Mathlin, Claudia Cooper, Lucinda Teoh, Naaheed Mukadam, Sube Banerjee, Yvonne Birks, Harriet Demnitz-King, Rachael Hunter

## Abstract

**Background:** People affected by dementia experience intersecting care inequalities. We explored relationships between ethnicity and health and social care resource use among people with dementia in an ethnically diverse urban region.

**Methods:** We conducted a retrospective observational cohort study using Discover-NOW, including patients with dementia between 1.4.2015 and 1.4.2025. We calculated ethnic density as the percentage of the Middle Layer Super Output Area (SOA) population self-identifying with the same ethnic group. Regression models, clustered by Local SOA, tested whether ethnic density moderated relationships between ethnicity and primary care, outpatient, inpatient, emergency and social care service use, controlling for sociodemographic characteristics, deprivation, comorbidities and time of diagnosis.

**Findings:** We included 30,704 people with dementia. People from Black and Mixed ethnic groups used more primary care, and those from Asian ethnic groups less primary and secondary care, than White ethnic groups. Rates of local authority social care packages were similar across ethnic groups. High ethnic density predicted fewer GP consultations in Black ethnic groups, but more in South Asian groups.

**Interpretation:** Among Black ethnic groups, primary care use was relatively high, especially in areas of low ethnic density, perhaps reflecting greater needs among communities at risk of racism and isolation. The trend towards increased primary care use among South Asian people in areas of higher ethnic density may reflect communities mitigating help-seeking hesitancy related to cultural and language barriers. Greater care integration could reduce care inequalities among minority ethnic communities who may experience fewer barriers to social relative to health care.

**Research in context:** 

**Evidence before this study:** We searched MEDLINE, EMBASE, and PsycINFO from inception to March 2026, using terms combining dementia, ethnicity, minority ethnic groups, health service use, social care, and ethnic density. We included studies in English reporting on health or social care utilisation among people with dementia from minority ethnic groups. Previous systematic reviews and cohort studies consistently documented that people from minority ethnic groups with dementia access planned health care, including primary care, memory services, and outpatient appointments, less frequently than White majority populations. A 2021 systematic review found that minority ethnic groups with dementia faced substantial barriers to accessing health services, including stigma, language barriers, unfamiliarity with services, and lack of culturally competent care. Studies using UK primary care data have similarly shown lower rates of physical health monitoring and psychotropic prescribing among minority ethnic groups with dementia. No prior study had examined how area-level ethnic density, the local concentration of co-ethnic residents, might moderate the relationship between ethnicity and care use in dementia, despite evidence from severe mental illness research that ethnic density is associated with lower mortality and may buffer against the health harms of racism and social isolation.

**Added value of this study:** This is the first study to examine whether ethnic density moderates the relationship between ethnicity and health and social care use among people with dementia. Using a large, linked, ethnically diverse primary care dataset of over 30,000 people with dementia registered at GPs in Northwest London, we found that people from Black and Mixed ethnic groups used primary care more frequently than White ethnic groups, while those from Asian ethnic groups used both primary and secondary care less. Strikingly, among Black ethnic groups, GP interaction rates were highest in areas of lowest ethnic density, consistent with greater unmet needs. Conversely, among South Asian groups, higher ethnic density was associated with a trend toward more frequent GP interactions, potentially reflecting greater cultural competency and community facilitation of help-seeking in these areas. We also provide, to our knowledge, the first quantitative evidence on social care package use by ethnicity in dementia, finding that over two-thirds of people with a dementia diagnosis received no community social care, with no statistically significant differences between ethnic groups.

**Implications of all the available evidence:** Persistent and patterned inequalities in health and social care access among people with dementia from minority ethnic groups, are shaped not only by individual and cultural factors but also by the ethnic composition of local communities. The finding that ethnic density modifies care-seeking in opposite directions for Black and South Asian groups underscores that minority ethnic communities are not homogeneous and that place-based strategies should be tailored to their needs. The England 10 Year Health Plan’s commitment to shifting care from hospital to community settings offers an opportunity to reduce these inequalities, but only if implementation is equitable. Neighbourhood care models that bring secondary care expertise into primary care settings could improve access for groups currently underusing specialist services. Greater integration of health and social care, with social care workers trained and resourced to play active roles in dementia management, may be particularly valuable for communities who appear more willing to engage with social than health care. Future research should examine ethnic density effects across other regions and healthcare systems, investigate the mechanisms linking community composition to care access, and evaluate whether integrated, culturally competent service models reduce the inequalities documented here.

## Introduction

Dementia is one of the most pressing global healthcare concerns; its prevalence is rising worldwide. The number of people with a dementia diagnosis in England increased from 427,176 in 2022 to 498,926 in 2025 but this represents only two-thirds of those with dementia; dementia is the UK’s leading cause of death (1). People with dementia have high care costs, with UK annual costs expected to reach £90 billion by 2040 (2).

*The 10 Year Health Plan for England* (3) declares government plans for paradigm shifts in service delivery, towards preventive, community care. Through the lens of dementia and frailty embodied in the plan’s commitment to a Modern Service Framework for Frailty and Dementia, central to realising this vision is the promise that proactive planned care management of dementia symptoms and comorbidities can prevent crises necessitating unplanned, emergency and hospital care, providing more acceptable and cost-effective care (4,5). If equitably implemented, this approach could increase care quality and reduce geographical care inequalities, as people with dementia who experience greater morbidity and mortality, who would benefit most, are over-represented in more deprived areas (6).

Current dementia care provision is not equitable. People from some minority ethnic groups receive less preventive care (7) and can be reluctant to use planned health (8) and social care services that are not perceived as culturally competent (9). These patterns of service use may help explain why minoritised ethnic status is associated with greater use of unplanned care (10).

To support equitable shifts towards community-based, preventive care, it is important to understand how geographical inequalities in health and care intersect with those related to ethnicity and culture. Among people with severe mental illness, ethnic density (the concentration of specific ethnic groups within an area) has been associated with lower mortality, likely reflecting protective effects on health outcomes and health-seeking behaviour, via increased social support, stronger social cohesion and reduced exposure to racism in those areas (11).

Such effects may also be relevant to dementia care inequalities, but to our knowledge this has not been explored. Potential drivers include, in areas of higher ethnic density, more community support, leading to lower formal care use, or more culturally competent services that are more acceptable and widely used. We explored how relationships between ethnicity and health and social care resource use varied by area-level ethnic density among people with dementia in an ethnically diverse urban region.

## Methods

### Study design

We conducted a retrospective observational cohort study using demographic, health and social care data for people with dementia living in Northwest London between 1^st^ April 2015 and 1^st^ April 2025.

### Data source

The Discover-NOW London TRE (Trusted Research Environment) is an anonymised linked dataset of over 2.8 million primary care registered individuals, linking social, community primary and secondary care service data (12). Discover-NOW have secured Health Research Authority approvals so researchers do not require further ethical approval to use the dataset for research studies approved by the North West London Data Access Committee (REC reference: 18/WM/0323; IRAS project ID: 253449). Data access was granted upon approval of a Data Access Request Form by this committee (approval number 337). The data were accessed through the Snowflake data warehouse using an Azure Machine Learning instance hosted within the Discover-NOW TRE and retrospectively analysed between November 2025 and March 2026 by GM, RH and LT.

### Study Population

We included patients aged 40 and over, with a dementia diagnosis recorded between 1^st^ April 2015 and 1^st^ April 2025, and at least six months of follow up data after dementia diagnosis. We restricted the sample to individuals with complete data on covariates (age at diagnosis, gender, care home status, household structure, and number of long-term conditions). We excluded patients with more than 15 long term conditions (as implausible data).

### Outcomes

To capture health and social care resource use, we recorded the total number of: (a) primary care interactions (in-person, telephone or home visits); those for whom more than one interaction a day with a GP was recorded were removed, as implausible data; (b) inpatient spells (excluding mental health admissions); (c) secondary care outpatient attendances (excluding mental health services); (d) emergency department attendances; and (e) social care packages (identified from a locally agreed dataset containing care package information relating to patients registered with a North West London General Practitioner (GP); we restricted this analysis to people who were not living in residential care. Supplement 1 provides further details for each variable.

### Exposures

Our main exposure was individual ethnicity, recorded in primary care records using self-reported UK Census categories. Ethnicity was classified in seventeen categories, which were further grouped into seven broader categories: Black (African, Caribbean, Any other Black background); South Asian (Bangladeshi, Indian, Pakistani, any other Asian background); Chinese; White (White UK, Any other White background, Irish), and mixed/multiple (Any other mixed background, White and Asian, White and Black African, White and Black Caribbean); any other ethnic background, and unknown (missing ethnicity).

### Area-level covariates

Ethnic density was calculated at the level of the Middle Layer Super Output Area (MSOA), following an established approach (13). The average population of an MSOA in London is 8,346 (14). Using data from the 2021 England and Wales Census (15), ethnic density was defined as the percentage of residents of an MSOA from a given ethnic group.

Individuals were assigned an ethnic density value corresponding to their own ethnic group within their MSOA of residence. Descriptive analyses used ethnic density based on the 17 census categories. For regression analyses, we categorised ethnic density in three groups (low, medium, and high) using equal-width cut-points derived from the observed range within each ethnic group. This within-group approach was adopted to account for the variation in ethnic density distributions across groups.

Socioeconomic deprivation was measured using the English Index of Multiple Deprivation (IMD). The IMD summarises individual and community characteristics within Lower layer Super Output Areas (LSOAs) of approximately 1500 people. Based on the IMD, LSOAs were divided into five quintiles, from the most affluent (quintile one) to the most deprived (quintile five) (16).

### Individual level covariates

Demographic covariates included: age at dementia diagnosis; sex (male/female); care home residence (yes/no); and household structure (living alone/not living alone).

Clinical covariates comprised a count of comorbid long-term conditions, derived from the 15 Quality Outcomes Framework chronic disease indicators (excluding dementia) recorded in the patient indicator table (17).

### Statistical Analysis

Annual observation periods were defined from the date of dementia diagnosis, with follow-up censored at the date of death or last recorded healthcare contact. Person-years of observation were calculated for each patient, and outcome counts were annualised and aggregated at the individual level for analysis. We summarised demographic characteristics of the sample and annual counts of health and social care use by ethnic groups.

Missing ethnicity, ethnic density and IMD were retained as a separate ‘Unknown’ category. For the remaining covariates, a complete-case analysis was undertaken. The only covariates with missing data were care home residence (27.9%) and household structure (28.3%) which were removed using listwise deletion. As the randomness of the missing data could not be determined, imputation was not used as it may have introduced bias (18). We used standard summary statistics to describe the data.

To examine the association between ethnicity and each annualised outcome, count regression models were fitted. Both Poisson and negative binomial regression models were evaluated based on model fit comparing AIC (Akaike Information Criteria) and BIC (Bayesian Information Criteria). Negative binomial regressions were selected for GP interactions due to over-dispersion. For inpatient spells, outpatient appointments and emergency department visits, we fitted zero-inflated negative binomial models to account for the high proportion of individuals in the sample with no events. A zero-inflated Poisson model provided the best fit for social care package use.

All models were adjusted for age at diagnosis, gender, care home residence, household structure, long term condition count and diagnosis year. Models were clustered at the LSOA level and included total person years as an offset. Models were fitted sequentially: model 1 included ethnicity; model 2 additionally adjusted for ethnic density and IMD; model 3 included an interaction between ethnicity and ethnic density. Model 3 was only fitted for the GP consultation rate, due to sample sizes for other variables below those recommended for interaction analyses (19). Individuals with missing ethnic density were removed from the interaction model, due to limited interpretability, and small counts influencing the regression model at the interaction level. Results were expressed as incidence rate ratios (IRRs) with 95% confidence intervals. All analyses were conducted in R (Version 4.4.1), with statistical significance set at p<0.05.

## Results

Table 1 displays the demographic characteristics of the full cohort of people with dementia (n=30,704). Table 2 describes ethnic densities across MSOAs. Median ethnic density varied substantially by group, ranging from 34.0% (IQR (Interquartile range) 23.8-43.6) for people from White British backgrounds (indicating that, on average, around a third of MSOA populations were White British); and 26.7% (IQR 18.3-39.5) for people of Indian ethnicity to, for people of mixed White and Black African ethnicity, a median ethnic density of 0.7% (IQR 0.6-1.1), indicating that on average this group lived in areas where less than one in a hundred people were from this ethnic group.

**Table 1.** Characteristics of included cohort of people with dementia.

|  | **N (%) / mean (standard deviation)** |
| --- | --- |
| Gender |  |
| Female | 17934 (58.41%) |
| Male | 12770 (41.59%) |
| Ethnicity (Detailed categories) |  |
| White British | 11070 (36.05%) |
| Any other White background | 4520 (14.72%) |
| White Irish | 1528 (4.98%) |
| White and Asian | 93 (0.30%) |
| White and Black African | 81 (0.26%) |
| White and Black Caribbean | 214 (0.70%) |
| Any other mixed background | 173 (0.56%) |
| Chinese | 166 (0.54%) |
| Pakistani | 659 (2.15%) |
| Any other Asian background | 1557 (5.07%) |
| African | 798 (2.60%) |
| Caribbean | 1612 (5.25%) |
| Any other Black background | 417 (1.36%) |
| Any other ethnic group | 956 (3.11%) |
| Missing | 2915 (9.49%) |
| Care home residence |  |
| No | 12736 (41.48%) |
| Yes | 9076 (29.56%) |
| Missing | 8892 (28.96%) |
| Household structure |  |
| Living alone | 5795 (18.87%) |
| Not living alone | 15909 (51.81%) |
| Missing | 9000 (29.31%) |
| Died during follow-up |  |
| No | 14689 (47.84%) |
| Yes | 16015 (52.16%) |
| IMD Quintile |  |
| 1 | 4447 (14.48%) |
| 2 | 11641 (37.91%) |
| 3 | 7123 (23.20%) |
| 4 | 4636 (15.10%) |
| 5 | 2675 (8.71%) |
| Missing | 182 (0.59%) |
| Number of long term conditions | 5.18 (2.33) |
| Age at diagnosis (years) | 81.05 (9.78) |
| Age at death (years) | 86.17 (7.73) |

Data are n (%) for categorical variables and mean (SD) for continuous variables. IMD = index of multiple deprivation.

**Table 2:**
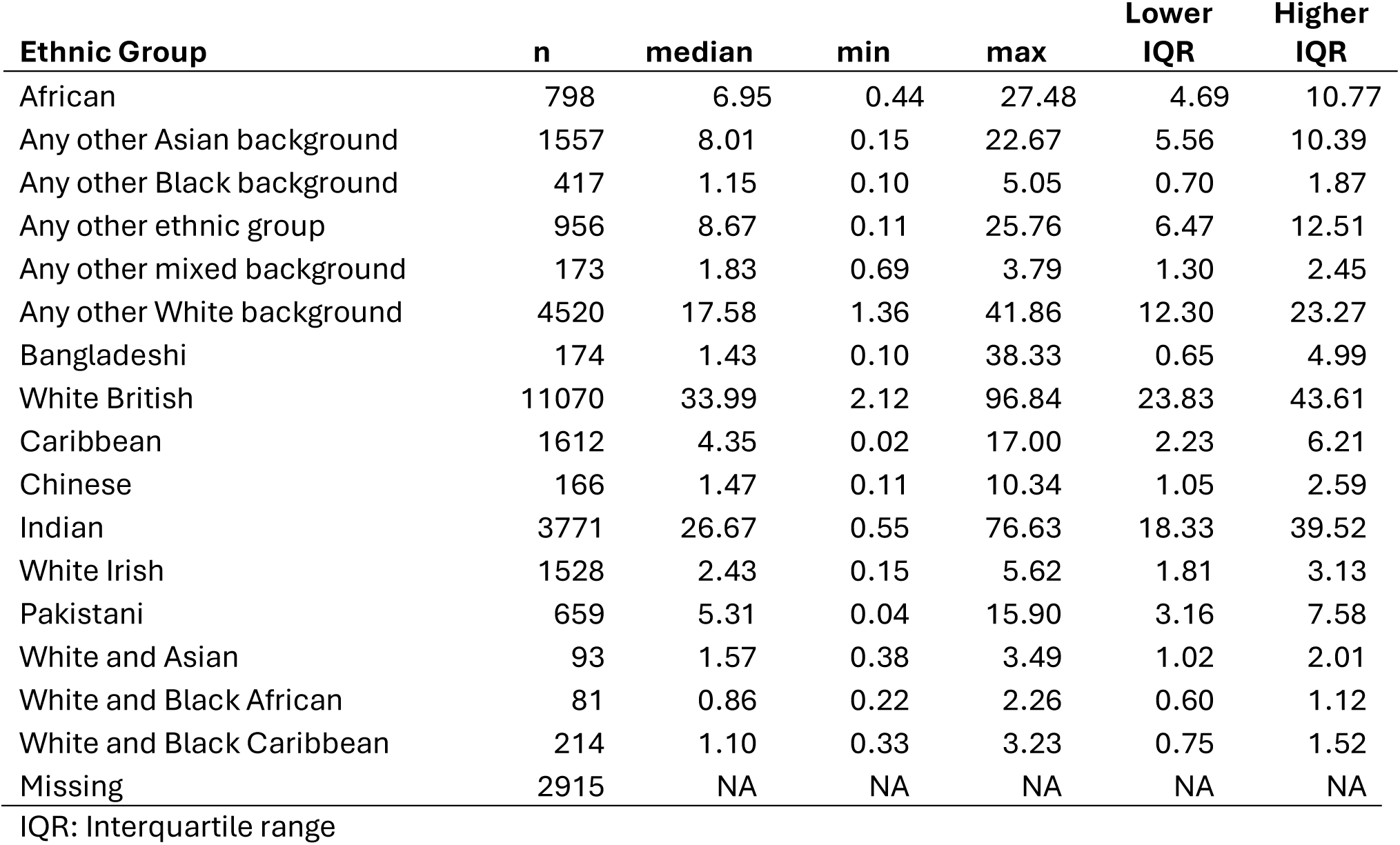
Ethnic density ranges by ethnic group.

### Health and social care use

Tables 3 and 4 describe the health and service use outcome mean rates per person/ year, by ethnic group (Table 3) and levels of ethnic density (Table 4). Figure 1 presents a flow diagram of the sample sizes for each outcome.

**Figure 1.**
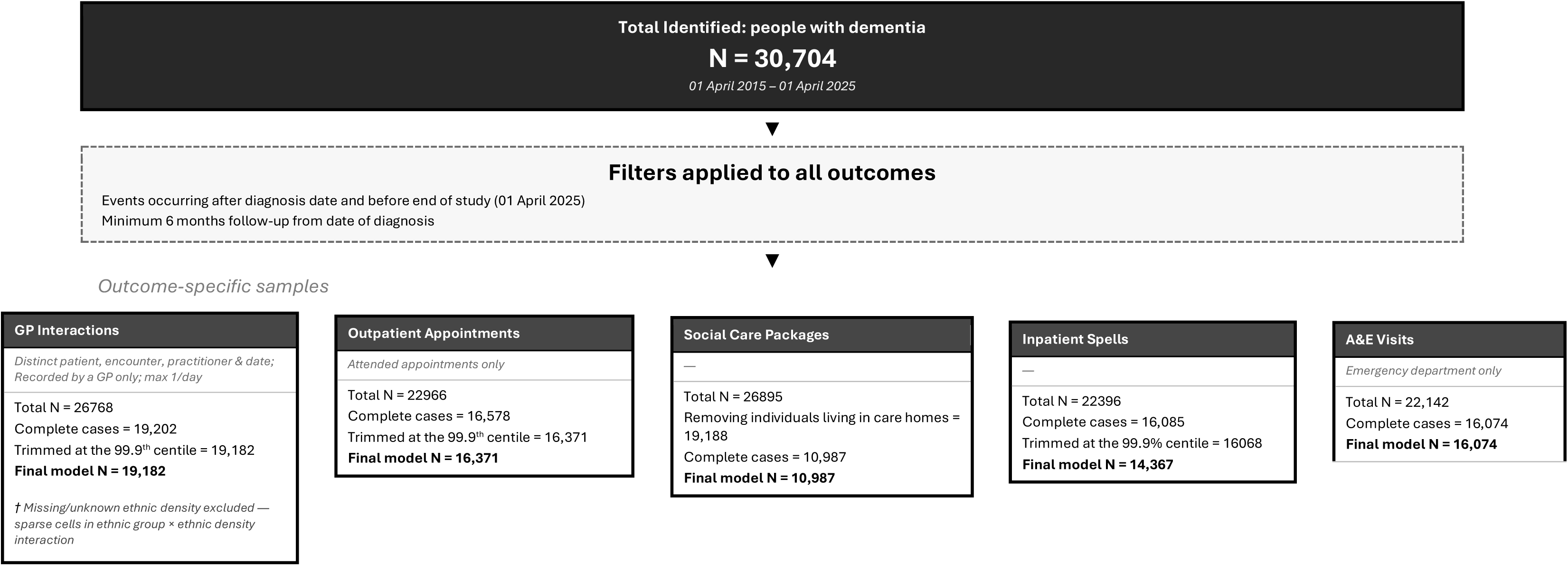
Participant Flow Diagram.

**Table 3.** Main health and social care use outcome average rates per person year.

(a) by ethnic group (n, mean rate, SD)
|  | White | Mixed/Multiple | Black | South Asian | Chinese/<br>Other Asian | Other | Missing |
| --- | --- | --- | --- | --- | --- | --- | --- |
| GP Interactions | 11035, 33.25 (31.04) | 353, 37.11 (45.84) | 1820, 36.22 (45.46) | 2633, 31.93 (29.96) | 990, 30.34 (25.98) | 592, 34.78 (38.20) | 1759, 31.36 (30.25) |
| Inpatient Spells | 9195, 1.21 (1.88) | 293, 1.23 (1.70) | 1522, 1.30 (2.11) | 2209, 1.26 (2.05) | 807, 1.08 (1.71) | 490, 1.17 (1.73) | 1535, 1.12 (1.72) |
| Outpatient Appointments | 9294, 4.05 (5.40) | 304, 4.25 (4.42) | 1570, 4.23 (4.90) | 2327, 4.69 (5.00) | 849, 4.14 (4.52) | 505, 4.36 (5.02) | 1522, 3.60 (4.62) |
| A&E Visits | 9261, 1.37 (1.90) | 288, 1.58 (2.39) | 1503, 1.32 (1.72) | 2201, 1.26 (1.72) | 810, 1.20 (1.54) | 479, 1.37 (1.90) | 1532, 1.27 (0.81) |
| Social Care Packages | 5721, 0.30 (0.52) | 204, 0.34 (0.55) | 1133, 0.43 (0.56) | 1875, 0.27 (0.48) | 682, 0.29 (0.48) | 392, 0.32 (0.51) | 971, 0.29 (0.50) |

(b) By ethnic density category (n, mean rate, SD)
|  | Low ethnic density | Medium ethnic density | High ethnic density | Missing |
| --- | --- | --- | --- | --- |
| GP Interactions | 8439, 32.86 (30.63) | 7862, 33.79 (32.31) | 1034, 33.52 (35.40) | 1847, 31.76 (30.63) |
| Inpatient Spells | 7115, 1.26 (2.12) | 6487, 1.19 (1.68) | 849, 1.17 (1.81) | 1600, 1.12 (1.71) |
| Outpatient appointments | 7205, 4.13 (5.24) | 6694, 4.24 (5.10) | 874, 4.23 (5.25) | 1598, 3.69 (4.98) |
| A&E Visits | 7163, 1.37 (1.89) | 6468, 1.33 (1.86) | 849, 1.24 (1.73) | 1594, 1.27 (1.81) |
| Social Care Packages | 4551, 0.32 (0.52) | 4750, 0.30 (0.52) | 656, 0.28 (0.49) | 1021, 0.29 (0.51) |

**Table 4.** Incident Rate Ratios (IRR) and 95% Confidence Intervals for relationships of health and social care use with ethnicity (model 1) and additionally controlling for area ethnic density and deprivation (model 2); p<0.05 indicated in bold.

| Model | Ethnic group | GP Interactions | Inpatient Spells | Outpatient Appointments | A&E Visits | Social Care Packages |
| --- | --- | --- | --- | --- | --- | --- |
| 1 | Black | **1.05 (1.01, 1.09)** | 1.04 (0.98, 1.11) | 0.97 (0.92, 1.02) | 0.98 (0.92, 1.04) | 1.04 (0.98, 1.10) |
|  | Chinese/Other Asian | **0.94 (0.90, 0.98)** | **0.86 (0.79, 0.94)** | **0.89 (0.83, 0.96)** | **0.89 (0.82, 0.97)** | 1.04 (0.96, 1.12) |
|  | Mixed/Multiple | **1.08 (1.02, 1.15)** | 1.02 (0.95, 1.09) | 0.99 (0.88, 1.11) | 1.08 (0.95, 1.23) | 1.04 (0.90, 1.20) |
|  | Other | 0.99 (0.95, 1.04) | **1.02 (0.89, 1.18)** | **0.88 (0.81, 0.97)** | 0.91 (0.82, 1.01) | 1.03 (0.94, 1.14) |
|  | South Asian | 0.99 (0.96, 1.01) | 0.89 (0.8, 1) | 0.96 (0.92, 1.01) | **0.88 (0.83, 0.94)** | 1.00 (0.94, 1.06) |
| 2 | Black | **1.04 (1.01, 1.08)** | 0.95 (0.89, 1.01) | 0.97 (0.92, 1.03) | 0.95 (0.89, 1.01) | 1.03 (0.96, 1.09) |
|  | Chinese/Other Asian | **0.94 (0.90, 0.97)** | 1.01 (0.95, 1.08) | **0.9 (0.83, 0.96)** | **0.87 (0.8, 0.95)** | 1.03 (0.95, 1.11) |
|  | Mixed/Multiple | **1.08 (1.01, 1.15)** | **0.84 (0.77, 0.92)** | 0.99 (0.89, 1.11) | 1.07 (0.94, 1.22) | 1.03 (0.89, 1.19) |
|  | Other | 0.99 (0.94, 1.04) | 0.69 (0.37, 1.29) | **0.89 (0.81, 0.97)** | 0.91 (0.82, 1.01) | 1.03 (0.94, 1.14) |
|  | South Asian | 0.98 (0.95, 1.01) | 0.95 (0.83, 1.1) | 0.96 (0.92, 1.01) | **0.87 (0.83, 0.93)** | 0.99 (0.93, 1.05) |

Note. White ethnicity is the reference group. All models controlled for age at diagnosis, gender, care home residence <sup>a</sup>, household structure, long term condition count, diagnosis year, clustered by LSOA and included total person years as the offset. Results are expressed as incidence rate ratios (IRRs) with 95% confidence intervals.
<sup>a</sup>The social care packages analysis did not include care home residence due to only including those not in a care home.

### GP Interactions

Six-month follow-up data regarding GP interactions were available for 26,768/30,704 (87.2%) of individuals. After removing individuals with missing care home or household group data (n=7,566), and interaction rates in the 99.9^th^ centile (n=20), 19,182 (62.5%) were included in the analytic cohort. All individuals had GP interactions during the study follow-up period; the median rate of interactions per person/year was 25.6, (IQR, 17.2 – 38.4). Analyses included a total of 65,536.5 person years. Participants were followed up for a mean of 3.9 (Standard Deviation, SD = 2.3) years.

Compared to people of White ethnicity (mean rate: 33.3, SD = 31.0), people with dementia from Black and Mixed/Multiple ethnic groups had significantly higher mean rates of GP consultations (37.1, SD=45.8), while those from Chinese/ Other Asian ethnic groups had significantly lower rates (30.3, SD=26.0) (Table 3). In adjusted analyses, people from Black (IRR (Incident Rate Ratios): 1.1, 95% CI: 1.0 -1.1) and Mixed/multiple ethnic groups (1.1, 1.0-1.2) consulted GPs more frequently, and those from Chinese/other Asian ethnic groups less frequently (0.9, 0.9-1.0), compared to people of White ethnicity (Table 4).

Table 5 shows how adding an interaction term between ethnic category and ethnic density moderated these findings. Rates of GP consultation were highest among Black people from areas of lowest ethnic density, compared to those living in more ethnically dense areas. While South Asian ethnic groups had similar rates of GP interactions to White ethnic groups, rates of GP consultation tended to be higher than White British groups in areas with high ethnic density (though this difference did not attain statistical significance).

**Table 5.** Incident Rate Ratios (IRR) and 95% Confidence Intervals for relationships of GP interactions with ethnicity (model 2) additionally adjusting for ethnic density by ethnicity interactions.

| Effects |  | GP Interactions |
| --- | --- | --- |
| Main | Black | **1.07 (1.03, 1.12)** |
|  | Chinese/Other Asian | 0.96 (0.90, 1.00) |
|  | Mixed/Multiple | 1.04 (0.95, 1.13) |
|  | Other | 1.00 (0.93, 1.08) |
|  | South Asian | 0.96 (0.93, 1.00) |
| Interaction ethnic density | Black x MED | **0.92 (0.85, 0.99)** |
|  | Chinese x MED | 0.98 (0.90, 1.07) |
|  | Mixed/multiple x MED | 1.05 (0.92, 1.20) |
|  | Other x MED | 0.98 (0.88, 1.10) |
|  | South Asian x MED | 1.04 (0.97, 1.11) |
|  | Black x HED | **0.76 (0.65, 0.89)** |
|  | Chinese/other x HED | 0.95 (0.77, 1.19) |
|  | Mixed/ multiple x HED | 1.23 (0.96, 1.57) |
|  | Other x HED | 0.97 (0.81, 1.16) |
|  | South Asian x HED | 1.14 (0.97, 1.34) |

### Outpatient appointments

Six months follow-up data were available for 22,966/30,704 (74.8%) of the main cohort (Figure 1). After removing individuals with missing care home or household group data (n=6578) and trimming data at the 99.9^th^ centile (n=17), the analytic cohort comprised16,371 individuals, of whom 13.4% (n = 2,195) had no outpatient appointments recorded during the study period. The median rate of outpatient appointments per person/ year was 2.7 (IQR = 1.0-5.5). The mean number of years of follow-up was 3.6 (SD = 2.3).

All minority ethnic groups had lower rates of outpatient attendances than White ethnic groups, after adjusting for covariates. Rates for those of Chinese and other Asian ethnicity, and Other ethnicity were 10.0% (95% CI: 0.8-1.0) and 9.1% (95% CI: 0.8-1.00) lower, respectively, than the White reference group in adjusted analyses (Table 4).

### Community social care packages

Six-month follow-up data were available for 26,895/30,704 of individuals in the main cohort. After removing individuals living in a care home (n = 7,707) and with missing household group data (n=8,210), 10,978 individuals remained, of whom 68.00% (7,470) did not have any social care packages during the study period.

The median rate of local authority community social care packages per person year was 0 (IQR = 0 – 0.6). The mean number of years of follow-up was 3.7 (SD = 2.0). There were no significant differences between ethnic groups in terms of the rate of use of community social care.

### Inpatient care

Six months follow-up data regarding inpatient spells were available for 22,396/30,704 (72.9%) of individuals; after removing individuals with missing care home or household group data (n=6,311) and trimming data at the 99.9^th^ centile (n=17), 16,068 individuals were included in this analysis, of whom 30% (n = 4,829) had no inpatient spells during the study period. The median rate of inpatient spells per person/ year was 0.7 (IQR = 0.0-1.6). The mean number of years of follow-up was 3.6 (SD = 2.2). Rates of inpatient admission were 6% lower among people from Mixed/multiple ethnic groups (IRR=0.8, 95% CI: 0.8-0.9), compared to White ethnic groups after controlling for variables studied (Table 4).

### Emergency department visits

Six months follow-up data regarding emergency department visits was available for 22,142/30,704 of individuals in the main cohort. After removing individuals with missing care home or household group data (n=6,037) and trimming data at the 99.9^th^ centile, 16,074 individuals were included in this analysis. 26.5% of individuals (4,254) had no emergency department visits during the study period. The median rate of visits per person/ year was 0.80 (IQR = 0.0-1.8). The mean number of years of follow-up was 3.7 (SD = 2.2). Relative to White ethnic groups, those of Chinese/other Asian (IRR=0.9, 95% CI: 0.8-1.0) and South Asian ethnicities (IRR=0.9, 95% CI: 0.8-0.9) had 13% lower rates of attendances in adjusted analyses (Table 4).

## Discussion

This is the first study to explore how ethnic density influences relationships between ethnicity and service use. We found important differences across ethnic groups compared with people of White ethnicity. People with dementia from Black and Mixed/multiple ethnic groups interacted with GPs more frequently, while those from Chinese/other Asian ethnic groups had lower rates of GP use and outpatient appointments. Emergency department visits were also lower among Chinese/other and South Asian, compared to White ethnic groups. Patterns varied by ethnic density. Among Black ethnic groups, GP interaction rates were higher in areas of low ethnic density, whereas among South Asian ethnic groups, GP consultation rates tended to be higher in areas of higher ethnic density.

Among Black ethnic groups, higher rates of GP use suggest that they may experience fewer access barriers to primary care, relative to outpatient care, which we would have expected to be accessed at similarly higher rates to reflect the greater level of need indicated by patterns of primary care use. Lower ethnic density was associated with more frequent GP interactions. Previous research has linked lower ethnic density with worse health outcomes, potentially due to greater exposure to racism and low community support (11). This may explain the pattern observed here, whereby higher use of GPs reflects greater underlying morbidity alongside potential barriers to accessing secondary care.

Across primary and secondary healthcare, the low rates of service use by people from Asian ethnic groups accord with findings from previous studies involving South Asian people with dementia (8,9). Reasons for low referral rates and access to secondary care among minority ethnic groups include unfamiliarity with services, economic barriers due to distance, complex booking systems inaccessible to non-English speakers; the need to be accompanied by a carer; and stigma associated with dementia (20). Interestingly, the low rates of primary care use were attenuated in areas of high ethnic density in South Asian ethnic groups. In such areas, services may be more likely to be culturally competent, for example, access to GPs from the same ethnic group and community support to translate and facilitate appointments.

To our knowledge, no study has explored how social care service use might vary between ethnic groups among people with dementia. The data records local authority community care package use, which is means tested. Perhaps the most striking finding is that over two-thirds of people with a dementia diagnosis did not receive any social care. This reflects findings from previous studies indicating that ethnic minority populations experience barriers to accessing social care (21). We did not find any differences in social care access between ethnic groups, though privately purchased social care was not included, so higher levels of deprivation among minority ethnic groups may have negatively confounded findings.

*The 10 Year Health Plan for England* (3) anticipates that proactive management of dementia symptoms and comorbidities in planned care encounters will prevent crises necessitating unplanned, emergency and hospital care. People with dementia from underserved groups experience more comorbidities (6) and lower access to care, so it is surprising that we did not find them to access more unplanned care. Perhaps the effect of having dementia (compared with not) on unplanned service use is greater than ethnic variation among those with dementia. Inequalities in outcomes may be driven by barriers to access to both planned and unplanned care. Person-centred, integrated care is predicated on the intention to provide care where people present, rather than where the system requires them to be. Neighbourhood care models enabling access to secondary care in primary care settings, could, based on our findings, improve access for people from minority ethnic groups, who appear to face barriers to accessing secondary care. Social care workers are also well placed to implement integrated dementia care models, and to promote health and wellbeing, though currently their agency to realise this potential to be fully active members of the multidisciplinary team is limited by poor access to training and supervision and resource constraints in the sector (22,23).

### Strengths and limitations

Previous studies reporting lower service use among people with dementia from minority ethnic groups, have not accounted for the interactive effects of community (7,24). To our knowledge, this is the first study to examine the interaction between ethnicity and ethnic density in relation to service use. We acknowledge important limitations.

While smaller than national datasets such as the Clinical Practice Research Datalink or The Health Improvement Network, the Discover-NOW TRE provides richer data linkage and is more ethnically diverse, while remaining broadly comparable to the UK population in terms of age, gender distribution, and chronic disease prevalence (25). Nonetheless, as the dataset only includes people from one London area, findings may not be immediately generalisable nationally, or internationally.

As with all routinely collected electronic health data, missing data are a key limitation. Ethnicity data may not be missing at random, for example if individuals concerned about discrimination are less willing to disclose it. We originally intended to include mental health outpatient and inpatient attendances, but high rates of missing data precluded this. Moreover, our sample was limited to individuals with a recorded dementia diagnosis, excluding the third of those with dementia who are undiagnosed. People from minority ethnic groups likely have higher rates of undiagnosed dementia (26), which may bias our findings. Those with a formal dementia diagnosis are more likely to be connected to mechanisms for social care so may have greater ability to access social care, especially if they have lower incomes; thus inequalities in social care access for minority groups may have been underestimated. Excluding people with missing data for living situation and care home residence may also bias findings.

Another limitation that arose from using electronic health data that are not designed specifically for research, is that certain variables, such as reason for GP visit were not consistently coded. Due to the breadth of GP read codes used we could not systematically remove administrative or preventive GP consultations which might explain why rates of GP use are higher than expected.

Even in this large dataset, we lacked power to examine interactions between ethnic group and density for outcomes beyond GP consultations. There are also limitations in the extent to which area-level variables apply at the individual level. IMD has been criticised as individuals may not share the deprivation level of the area in which they live (27). Ethnic categories encompass heterogeneous populations, with diverse migration histories, cultural and linguistic backgrounds and socioeconomic statuses, so only approximate individual-level exposure (28).

### Conclusions

People from minority ethnic groups used less secondary health care than people from White ethnic groups. People from Black and Mixed ethnic groups interacted with GPs more, while those from Chinese and other Asian groups have fewer GP interactions. In people from Black ethnicity, those in areas of lower ethnic density had more GP interactions, while in South Asian ethnic groups, living in a higher ethnic density area predicted a higher GP interaction rate. These patterns may reflect the dual influence of community: supporting better health outcomes and help-seeking behaviours. Minority ethnic groups tended to access secondary care less, highlighting the potential value of greater health and social care integration in reducing inequalities in access to appropriate services.

## Data Availability Statement

The data used in this paper cannot be shared directly. Discover-NOW welcome applications to use their data on a project-specific basis, with data access subject to their approvals processes and relevant data sharing agreements, described on their respective websites.

## Acknowledgments

This research is funded through the NIHR Policy Research Unit in Dementia and Neurodegeneration – Queen Mary University of London (reference NIHR206110). The views expressed are those of the author(s) and not necessarily those of the NIHR or the Department of Health and Social Care.

## Supplementary Information – Outcome Identification

For all outcomes, events had to occur after the diagnosis date and before end of study (01 April 2025) and each person had to have a minimum of 6 months follow-up data from date of diagnosis.

- GP interactions were identified from the ‘DWH.FACT_COMPASS_OBSERVATION_DID’ table, identifying distinct patient, encounter, practitioner & date entries, recorded by a GP only and limited to a maximum of 1 interaction per day.
- Inpatient care spells were identified from the ‘DWH.FACT_SUS_APC_SPELL’ table and all non-mental health related admitted patient care spells were identified.
- Outpatient appointments were identified using the ‘DWH.FACT_SUS_OUTPATIENT’ table and filtered to only include appointments recorded as attended.
- A&E visits were identified from the ‘DWH.FACT_SUS_AE_DID’ table with only visits to an emergency department (no urgent treatment centre admissions included).
- Social care packages were identified though the ‘DWH.FACT_SOCIAL_CARE_B13B_DID’ table through counting the number of times a date of a social care package occurring was recorded.

**Supplementary Table 1.** Incident Rate Ratios (IRR) and 95% Confidence Intervals for relationships of health and social care use and all covariates (model 1) and additionally controlling for area ethnic density and deprivation (model 2); p<0.05 indicated in bold.

| Model |  | GP Interactions | Inpatient Spells | Outpatient Appointments | A&E Visits | Social Care Packages |
| --- | --- | --- | --- | --- | --- | --- |
| 1 | Black | 1.05 (1.02, 1.09) | 1.04 (0.98, 1.11) | 0.97 (0.92, 1.02) | 0.98 (0.92, 1.04) | 1.04 (0.98, 1.1) |
|  | Chinese/Other Asian | 0.94 (0.9, 0.98) | **0.86 (0.79, 0.94)** | **0.89 (0.83, 0.96)** | **0.89 (0.82, 0.97)** | 1.04 (0.96, 1.12) |
|  | Mixed/Multiple | 1.08 (1.02, 1.15) | 1.02 (0.89, 1.18) | 0.99 (0.88, 1.11) | 1.08 (0.95, 1.23) | 1.04 (0.9, 1.2) |
|  | Other | 0.99 (0.95, 1.04) | **0.89 (0.8, 1)** | **0.88 (0.81, 0.97)** | **0.91 (0.82, 1.01)** | 1.03 (0.94, 1.14) |
|  | South Asian | 0.99 (0.96, 1.01) | 0.95 (0.89, 1.01) | 0.96 (0.92, 1.01) | **0.88 (0.83, 0.94)** | 1 (0.94, 1.06) |
|  | Age at diagnosis | **1 (1, 1)** | 1 (1, 1) | **0.98 (0.98, 0.99)** | **1 (1, 1)** | **1 (1, 1.01)** |
|  | Gender (Male) | **1.07 (1.05, 1.09)** | **1.3 (1.25, 1.35)** | **1.2 (1.16, 1.24)** | **1.25 (1.2, 1.3)** | 1 (0.97, 1.04) |
|  | Living in a care home (True) | **1.08 (1.05, 1.11)** | 1.02 (0.96, 1.07) | 0.71 (0.68, 0.75) | **1.17 (1.12, 1.23)** | - |
|  | Household structure (Living alone) | **1.07 (1.05, 1.1)** | **1.06 (1, 1.11)** | **0.94 (0.9, 0.98)** | **1.14 (1.08, 1.2)** | **1.1 (1.06, 1.15)** |
|  | Long term condition count | **1.09 (1.08, 1.09)** | **1.13 (1.13, 1.14)** | **1.08 (1.08, 1.09)** | **1.11 (1.1, 1.12)** | 1.01 (1, 1.01) |
|  | diagnosis year 2016 | 1.04 (1, 1.09) | **1.32 (1.21, 1.42)** | **1.55 (1.45, 1.66)** | **1.31 (1.21, 1.41)** | 0.96 (0.89, 1.04) |
|  | diagnosis year 2017 | **1.14 (1.09, 1.19)** | **1.97 (1.81, 2.13)** | **2.43 (2.26, 2.61)** | **2.09 (1.94, 2.26)** | 1.07 (0.99, 1.15) |
|  | diagnosis year 2018 | **1.16 (1.11, 1.21)** | **1.93 (1.77, 2.09)** | **1.94 (1.81, 2.08)** | **2.04 (1.89, 2.2)** | 1.07 (1, 1.16) |
|  | diagnosis year 2019 | **1.25 (1.2, 1.3)** | **1.97 (1.81, 2.14)** | **1.75 (1.63, 1.88)** | **2.05 (1.9, 2.22)** | **1.11 (1.03, 1.2)** |
|  | diagnosis year 2020 | **1.43 (1.37, 1.49)** | **2.11 (1.94, 2.3)** | **1.87 (1.74, 2.01)** | **2.24 (2.07, 2.43)** | **1.21 (1.12, 1.31)** |
|  | diagnosis year 2021 | **1.44 (1.38, 1.5)** | **2.18 (2, 2.37)** | **2.09 (1.95, 2.24)** | **2.25 (2.08, 2.44)** | **1.27 (1.18, 1.38)** |
|  | diagnosis year 2022 | **1.42 (1.36, 1.48)** | **2.32 (2.13, 2.52)** | **2.04 (1.9, 2.19)** | **2.35 (2.17, 2.54)** | **1.26 (1.15, 1.37)** |
|  | diagnosis year 2023 | **1.52 (1.45, 1.58)** | **2.7 (2.46, 2.96)** | **2.37 (2.2, 2.55)** | **2.82 (2.58, 3.08)** | **1.29 (1.17, 1.42)** |
|  | diagnosis year 2024 | **1.64 (1.57, 1.71)** | **2.89 (2.56, 3.28)** | **2.74 (2.51, 2.99)** | **3.16 (2.81, 3.55)** | **0.82 (0.7, 0.95)** |
| 2 | Black | **1.04 (1.01, 1.08)** | 1.01 (0.95, 1.08) | 0.97 (0.92, 1.03) | 0.95 (0.89, 1.01) | 1.03 (0.96, 1.09) |
|  | Chinese/Other Asian | **0.94 (0.9, 0.97)** | **0.84 (0.77, 0.92)** | **0.9 (0.83, 0.96)** | **0.87 (0.8, 0.95)** | 1.03 (0.95, 1.11) |
|  | Mixed/Multiple | **1.08 (1.01, 1.15)** | 0.95 (0.83, 1.1) | 0.99 (0.89, 1.11) | 1.07 (0.94, 1.22) | 1.03 (0.89, 1.19) |
|  | Other | 0.99 (0.94, 1.04) | **0.87 (0.78, 0.97)** | **0.89 (0.81, 0.97)** | 0.91 (0.82, 1.01) | 1.03 (0.94, 1.14) |
|  | South Asian | 0.98 (0.95, 1.01) | 0.94 (0.89, 1) | 0.96 (0.92, 1.01) | **0.87 (0.83, 0.93)** | 0.99 (0.93, 1.05) |
|  | Ethnic density category (Medium) | 0.98 (0.96, 1) | 0.97 (0.93, 1.01) | 1 (0.96, 1.04) | 0.98 (0.94, 1.02) | 1 (0.95, 1.04) |
|  | Ethnic density category (High) | 1.02 (0.98, 1.07) | 0.98 (0.89, 1.07) | 0.99 (0.91, 1.06) | 0.96 (0.87, 1.05) | 1 (0.91, 1.1) |
|  | IMD quintile 2 | 0.96 (0.9, 1.02) | 0.95 (0.88, 1.02) | 1.01 (0.95, 1.07) | 0.98 (0.91, 1.05) | 1.06 (0.99, 1.14) |
|  | IMD quintile 3 | 0.95 (0.89, 1.01) | **0.92 (0.84, 1)** | 1.04 (0.97, 1.11) | **0.89 (0.82, 0.96)** | 0.98 (0.9, 1.06) |
|  | IMD quintile 4 | 0.95 (0.88, 1.02) | **0.83 (0.76, 0.91)** | 1.06 (0.99, 1.14) | **0.84 (0.77, 0.91)** | 0.98 (0.89, 1.08) |
|  | IMD quintile 5 | **0.83 (0.77, 0.9)** | **0.75 (0.67, 0.84)** | 1.02 (0.93, 1.11) | **0.73 (0.66, 0.81)** | **0.71 (0.6, 0.84)** |
|  | Age at diagnosis | 1 (1, 1) | 1 (1, 1) | **0.98 (0.98, 0.98)** | **1 (1, 1.01)** | **1 (1, 1.01)** |
|  | Gender (Male) | 1.07 (1.05, 1.09) | **1.3 (1.25, 1.35)** | **1.2 (1.16, 1.24)** | **1.24 (1.19, 1.28)** | 1 (0.96, 1.04) |
|  | Living in a care home (True) | 1.08 (1.05, 1.11) | 1.02 (0.97, 1.08) | **0.71 (0.68, 0.75)** | **1.18 (1.12, 1.24)** | - |
|  | Household structure (Living alone) | 1.07 (1.05, 1.1) | **1.06 (1, 1.11)** | **0.94 (0.9, 0.98)** | **1.14 (1.09, 1.2)** | **1.11 (1.06, 1.15)** |
|  | Long term condition count | 1.09 (1.08, 1.09) | **1.13 (1.12, 1.14)** | **1.08 (1.08, 1.09)** | **1.11 (1.1, 1.12)** | 1.01 (1, 1.01) |
|  | diagnosis year 2016 | 1.04 (1, 1.08) | **1.3 (1.2, 1.4)** | **1.55 (1.45, 1.66)** | **1.29 (1.2, 1.39)** | 0.97 (0.9, 1.04) |
|  | diagnosis year 2017 | **1.14 (1.09, 1.19)** | **1.94 (1.79, 2.11)** | **2.43 (2.26, 2.6)** | **2.05 (1.9, 2.21)** | 1.07 (0.99, 1.15) |
|  | diagnosis year 2018 | **1.16 (1.11, 1.21)** | **1.93 (1.78, 2.09)** | **1.94 (1.8, 2.08)** | **2.02 (1.87, 2.18)** | **1.08 (1, 1.16)** |
|  | diagnosis year 2019 | **1.25 (1.2, 1.3)** | **1.96 (1.8, 2.13)** | **1.75 (1.63, 1.87)** | **2.03 (1.88, 2.2)** | **1.11 (1.03, 1.2)** |
|  | diagnosis year 2020 | **1.43 (1.37, 1.49)** | **2.11 (1.94, 2.3)** | **1.87 (1.74, 2.01)** | **2.19 (2.02, 2.38)** | **1.21 (1.12, 1.31)** |
|  | diagnosis year 2021 | **1.44 (1.38, 1.5)** | **2.16 (1.99, 2.35)** | **2.08 (1.94, 2.23)** | **2.24 (2.07, 2.42)** | **1.28 (1.18, 1.38)** |
|  | diagnosis year 2022 | **1.42 (1.36, 1.48)** | **2.31 (2.13, 2.52)** | **2.03 (1.9, 2.18)** | **2.34 (2.16, 2.54)** | **1.25 (1.15, 1.37)** |
|  | diagnosis_year2023 | **1.51 (1.45, 1.58)** | **2.7 (2.47, 2.96)** | **2.37 (2.2, 2.55)** | **2.81 (2.58, 3.07)** | **1.29 (1.17, 1.42)** |
|  | diagnosis year 2024 | **1.64 (1.57, 1.72)** | **2.89 (2.56, 3.27)** | **2.74 (2.51, 2.99)** | **3.16 (2.82, 3.55)** | **0.82 (0.71, 0.96)** |

Note. White ethnicity, female gender, not living in a care home, not living alone, diagnosis year 2015 are the reference groups. All models were clustered by LSOA and included total person years as the offset. Results are expressed as incidence rate ratios (IRRs) with 95% confidence intervals.
<sup>a</sup>The social care packages analysis did not include care home residence due to only including those not in a care home.

**Supplementary Table 2.** Incident Rate Ratios (IRR) and 95% Confidence Intervals for relationships of GP interactions with all covariates (model 2) additionally adjusting for ethnic density by ethnicity interactions.

| Effects |  |  |
| --- | --- | --- |
| Main | Black | **1.07 (1.03, 1.12)** |
|  | Chinese/Other Asian | 0.95 (0.9, 1) |
|  | Mixed/Multiple | 1.04 (0.95, 1.13) |
|  | Other | 1 (0.93, 1.08) |
|  | South Asian | 0.96 (0.93, 1) |
|  | Ethnic density category (Medium) | 0.99 (0.96, 1.02) |
|  | Ethnic density category (High) | 1.04 (0.98, 1.1) |
|  | IMD quintile 2 | 0.94 (0.89, 1) |
|  | IMD quintile 3 | **0.93 (0.87, 1)** |
|  | IMD quintile 4 | **0.93 (0.87, 1)** |
|  | IMD quintile 5 | **0.82 (0.76, 0.89)** |
|  | Age at diagnosis | **1 (1, 1)** |
|  | Gender (Male) | **1.07 (1.05, 1.09)** |
|  | Living in a care home (True) | **1.08 (1.05, 1.11)** |
|  | Household structure (Living alone) | **1.07 (1.05, 1.1)** |
|  | Long term condition count | **1.09 (1.08, 1.09)** |
|  | diagnosis year 2016 | **1.04 (1, 1.08)** |
|  | diagnosis year 2017 | **1.14 (1.09, 1.19)** |
|  | diagnosis year 2018 | **1.16 (1.11, 1.21)** |
|  | diagnosis year 2019 | **1.25 (1.2, 1.3)** |
|  | diagnosis year 2020 | **1.43 (1.37, 1.49)** |
|  | diagnosis year 2021 | **1.44 (1.38, 1.5)** |
|  | diagnosis year 2022 | **1.42 (1.36, 1.48)** |
|  | diagnosis_year2023 | **1.51 (1.45, 1.58)** |
|  | diagnosis year 2024 | **1.64 (1.57, 1.72)** |
| Interaction | Black x MED | **0.92 (0.85, 0.99)** |
|  | Chinese x MED | 0.98 (0.9, 1.07) |
|  | Mixed/multiple x MED | 1.05 (0.92, 1.2) |
|  | Other x MED | 0.98 (0.88, 1.1) |
|  | South Asian x MED | 1.04 (0.97, 1.11) |
|  | Black x HED | **0.76 (0.65, 0.89)** |
|  | Chinese/other x HED | 0.95 (0.77, 1.19) |
|  | Mixed/ multiple x HED | 1.23 (0.96, 1.57) |
|  | Other x HED | 0.97 (0.81, 1.16) |
|  | South Asian x HED | 1.14 (0.97, 1.34) |

Note. White ethnicity, female gender, not living in a care home, not living alone, diagnosis year 2015 are the reference groups. All models were clustered by LSOA and included total person years as the offset. Results are expressed as incidence rate ratios (IRRs) with 95% confidence intervals.
<sup>a</sup>The social care packages analysis did not include care home residence due to only including those not in a care home.
MED= Medium ethnic density; HED = High ethnic density

## Notes

### Competing Interest Statement

The authors have declared no competing interest.

### Author Declarations

North West London Data Access Committee (reference: 18/WM/0323) and NHS IRAS project ID: 253449 gave ethical approval for this work.

